# Expansion of a low-cost, saliva-based PCR test for the detection of mpox virus

**DOI:** 10.1101/2023.06.12.23291295

**Authors:** Russell J Thomas, Orchid M. Allicock, Devyn Yolda-Carr, Katherine Fajardo, Sydney A. Steel, Chessley B. Blacklock, Theresa Zepeda, Maurice Brownlee, Shyam Saladi, James Parkin, Anne L. Wyllie

## Abstract

**Background:** Current recommendations for the diagnosis of mpox rely on lesion-swabs as the gold-standard specimen type, even though many patients experience symptoms prior to lesion-onset. Alternative sample types, such as saliva, which enable earlier detection could bolster the mpox response by mitigating transmission and facilitating access to antiviral treatments.

**Methods:** We evaluated five PCR assays and compared their detection of mpox DNA extracted from 30 saliva specimens collected in Spectrum SDNA-1000 tubes. We sequenced seven mpox-positive samples and assessed concordance with the primers and probes of the PCR assays. Following, we incorporated these PCR assays into a simplified, extraction-free protocol to evaluate its feasibility for testing raw (unsupplemented) saliva samples. To further explore the potential of this approach, we investigated the stability of mpox detection in raw saliva diluted 1:10 and 1:100 in mpox-negative saliva, after storage at 4°C, room temperature (∼19°C), 30°C, and 40°C for 72 hours and through simulated shipping conditions.

**Results:** Despite identifying three nucleotide substitutions in the CDC’s Monkeypox virus Generic Real-Time PCR Test’s primer sequences, we observed no difference in the mean Ct-values generated between assays. We successfully incorporated each assay into our saliva-based extraction-free PCR protocol. Detection in raw saliva following storage at 4°C, ∼19°C, and 30°C remained relatively stable for 24-48 hours and following simulated shipping conditions.

**Conclusions:** This pilot investigation supports a flexible, saliva-based, extraction-free PCR test as a promising approach for diagnosis, outbreak response or ongoing surveillance of mpox. With detection in raw saliva remaining stable for 24-48 hours and through simulated shipping temperatures, saliva-based sampling and simplified testing could reduce diagnostic costs, increase access to testing and address hurdles in low- and middle-income countries.

## BACKGROUND

Countries in Western and Central Africa have long dealt with outbreaks of mpox virus, with a likely rodent reservoir contributing to spillover [1]. Few outbreaks of mpox virus have occurred outside of Africa, and when they have, they have been largely self-limiting. However, in 2022, the virus spread globally, particularly in the sexual networks of men who have sex with men [2]. Much like the challenges seen with SARS-CoV-2, public health systems struggled as case counts rapidly grew, with testing bottlenecks and insufficient access to treatment and prophylaxis [2].

Mpox infection typically presents with a viral prodrome followed by the characteristic “pox” lesions. During the 2022 outbreak, studies highlighted that cases were experiencing oral or mucosal lesions, with some reporting oropharyngeal symptoms first [2,3]. These reports corroborate historical data; a study from the Democratic Republic of Congo found that 29% of cases had mouth/throat lesions, with 78% of cases also reporting a sore throat, with blood and pharyngeal samples confirming infection earlier than lesion swabs [4].

Through 2022, saliva emerged as a viable diagnostic specimen for mpox, demonstrating high sensitivity [5–9]. Notably, Allen-Blitz *et al*. found that testing saliva accurately identified 22 cases, four of which did not have a rash and one had no symptoms at all [5]. Hernaez *et al*. found systemic symptoms and lesions associated with higher viral loads in saliva and isolated infectious virus from 22/33 saliva samples [8]. When followed over the course of the infection, mpox has been detected in saliva at lower qPCR cycle threshold (Ct) values than from skin lesions [6,7] and oral swab samples [9]. In a systematic review, saliva, anorectal and skin lesion samples had the highest viral loads, all greater than that of pharyngeal samples [10]. Moreover, viral loads peaked earliest in saliva, within four days of symptom onset [10], with Brosius *et al*. detecting DNA and replication-competent mpox virus up to four days before symptom onset [11]. While detection of mpox in saliva typically declines within 14 days of symptoms onset [10], mpox virus has been reported in saliva 76 days after diagnosis [12]. The diagnostic implications for this suggest that mpox virus can circulate systemically prior to lesions and in some cases following their resolution, which raises questions surrounding the potential for asymptomatic transmission and opportunities for improved screening measures, particularly of close-contacts.

Together with saliva being a CDC-recognized source of transmission [13], the growing body of literature underscores the potential of saliva as a specimen type for the detection of mpox virus – its advantages including its ease of self-collection and ability to detect infection prior to lesion onset. With these benefits supporting serial sample collection from exposed individuals, this could identify cases earlier than lesion development. Early detection facilitates faster access to antivirals, which may lessen the severity of disease and concomitantly, facilitates earlier isolation, thus aiding public health workers in halting transmission chains. Therefore, in this study, we evaluated the potential of a simplified PCR test for the detection of mpox virus in saliva, aiming to enhance outbreak response and sustainable surveillance efforts.

## METHODS

### Ethics statement

This study was conducted with Institutional Review Board approval from Yale Human Research Protection Program (Protocol ID. 2000033293), which allowed for the use of remnant clinical samples and was considered as non-human subjects research. No personal identifiable information was permitted for use in this study.

### Samples

We received 30 known mpox-positive saliva samples in Spectrum SDNA-1000 collection devices from FlowHealth (Culver City, CA). These collection devices contain patented preservative media to stabilize analytes. Additionally, we received five raw (unsupplemented) mpox-positive saliva samples from Neelyx Labs (Wood Dale, IL) that were collected from patients with positive lesion-swabs.

### PCR assay performance

Using synthetic mpox virus DNA (ATCC VR-3270SD, ATCC Manassas, VA), we assessed the limit of detection (LOD) for five different PCR assays: the Logix Smart™ Mpox (2-Gene) RUO (Co-Diagnostics, Inc., Salt Lake City, UT), the Mirimus MPOX RT-PCR assay (Mirimus Labs, Brooklyn, NY), the assay targeting Clade II developed by Yu Li *et al*. [14], and the CDC assays consisting of targets for both mpox virus and non-variola orthopoxvirus [15] (**Table 1**). As the sequences for the primers and probes of the Logix Smart™ Mpox (2-Gene) RUO assay from Co-Diagnostics, Inc. are proprietary, they are not listed. Two target assays had been recommended by the US Food and Drug Administration (FDA) and supported by assay developers in order to provide two, independent outputs to corroborate each other, and to make the assay more robust against future variants that could emerge over time.

**Table 1.**
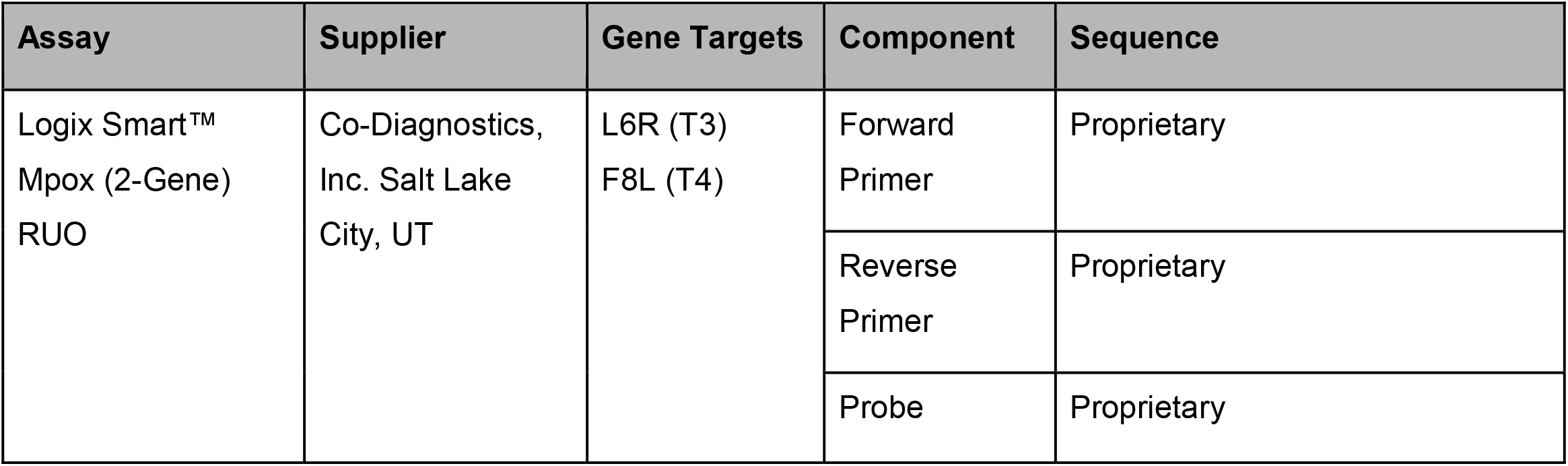
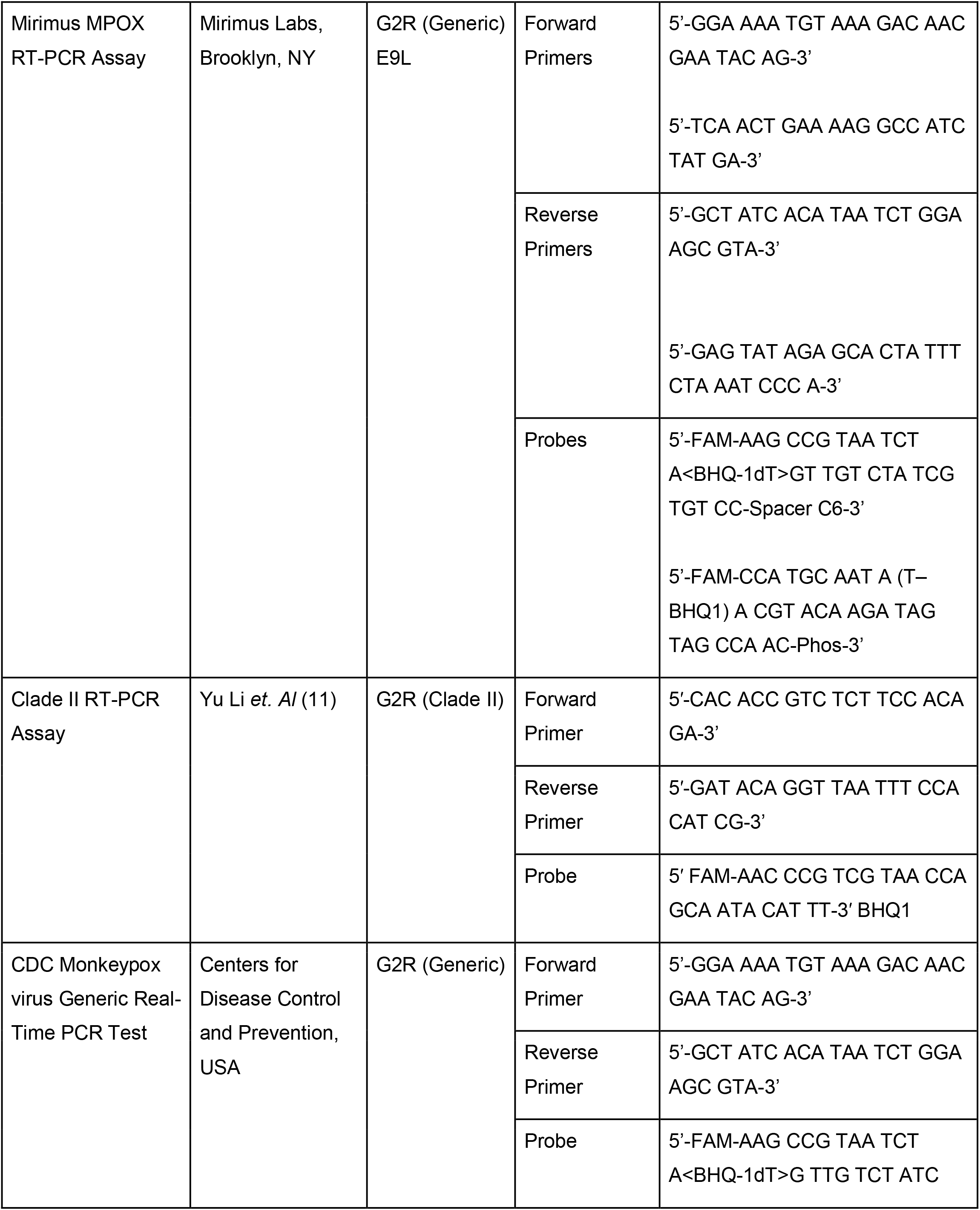
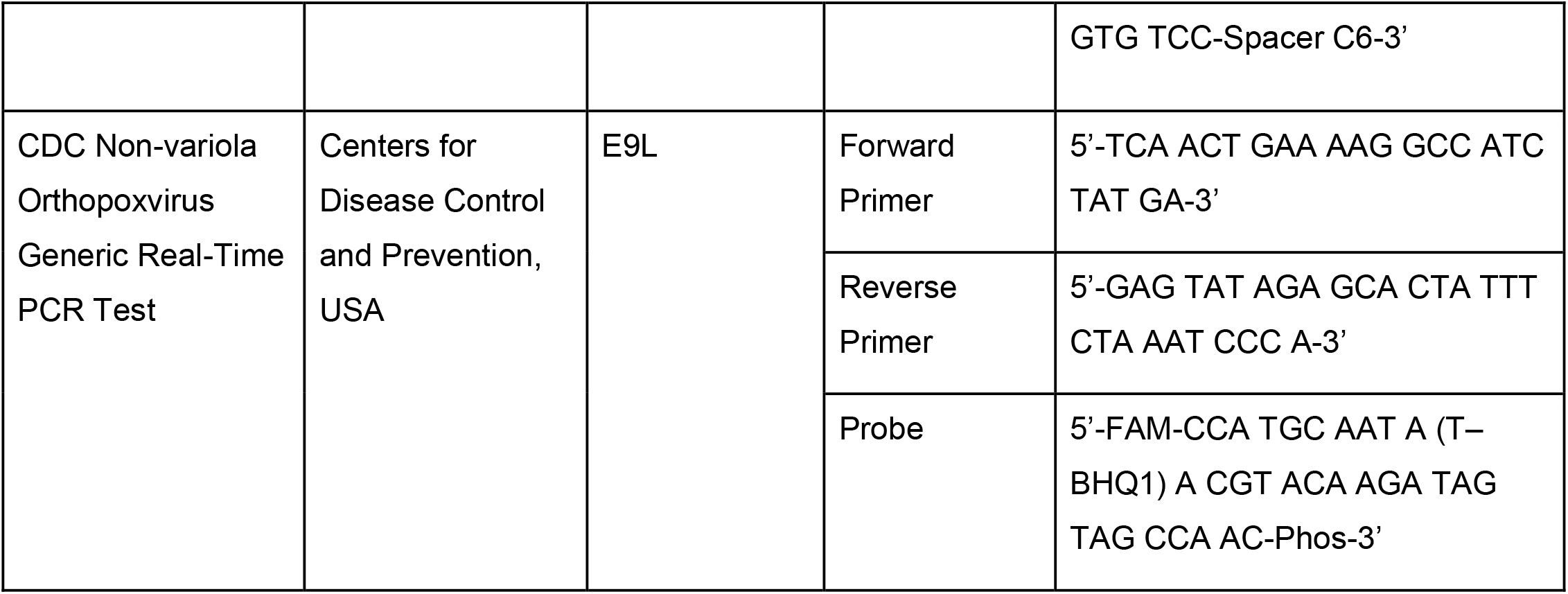
Gene targets, primers, and probe sequences of the mpox PCR assays evaluated.

Triplicate tests were performed using two-fold dilution series of the synthetic DNA controls in water, ranging from 100 copies/µl to 1 copy/µl. Reaction mixes, including primer and probe concentrations, either followed the manufacturer’s instructions (Co-Diagnostics), the CDC guidance for the detection of non-variola orthopoxvirus (CDC and Clade II assays) [15], or as detailed in **Supplementary Table 1** (Mirimus). Assays other than the Logix Smart™ Mpox (2-Gene) RUO assay from Co-Diagnostics were run with the Luna® Universal One-Step RT-qPCR Kit (New England Biolabs, MA). Thermocycler conditions for each assay are provided in **Supplementary Table 2**. For the Mirimus MPOX RT-PCR assay, the CDC Monkeypox virus Generic Real-Time PCR Test, and the CDC Non-variola Orthopoxvirus Real-Time PCR Test, samples were considered positive when Ct values <40. For the Logix Smart™ Mpox (2-Gene) RUO assay and the assay targeting Clade II samples were considered positive when Ct values were <42.

We then validated the five PCR assays on the 30 samples received from FlowHealth. In each assay, 5 µl of DNA template, extracted using the MagMAX™ Viral/Pathogen Nucleic Acid Isolation Kit (ThermoFisher Scientific, MA), was tested in a 20 µl reaction volume.

### Extraction-free workflows

To explore the potential of streamlined, extraction-free PCR testing of saliva for the detection of mpox virus, we validated each of the three SalivaDirect workflows developed for the detection of SARS-CoV-2 [16] on three raw saliva samples from Neelyx Labs. Briefly, workflow one includes the addition of proteinase K followed by heat inactivation at 95°C for 5 minutes, workflow two includes heat treatment at 95°C without addition of proteinase K, and workflow three includes heat pre-treatment at 95°C for 30 minutes, followed by addition of proteinase K, then heat inactivation at 95°C for 5 minutes.

### Stability of detection of mpox virus in raw, unsupplemented saliva

Having demonstrated the stability of detection of other respiratory pathogens in raw saliva [17–19], we explored the stability of the detection of mpox virus DNA in the raw mpox-positive samples diluted 1:10 and 1:100 into mpox-negative saliva. Saliva lysates prepared from the 1:10 dilutions using workflow one of the extraction-free protocol were tested with the Logix Smart™ Mpox (2-Gene) RUO assay and the lysates prepared from the 1:100 dilutions were tested with the Mirimus MPOX RT-PCR assay. Time zero values were adjusted based on this dilution factor, for which a dilution of 1:10 would yield approximately a +3 increase in Ct value. To test stability, we assessed the change in Ct values at 24, 48, and 72 hours of incubation at 4°C, room temperature (∼19°C), 30°C, and 40°C. Due to limited volume of raw saliva, we further assessed the stability of detection using the DNA extracted from a subset of the saliva samples in Spectrum SDNA-1000 collection devices, spiked into raw mpox-negative saliva at a ratio of 1:100, and incubated at room temperature for up to 72 hours.

Additionally, we assessed the stability of mpox detection following incubation through a modification of the US FDA’s summer and winter shipping profile conditions, modeled after International Safe Transit Association (ISTA) 7D shipping standards. Due to limited sample volume, only three samples were run through the simulated shipping profiles, and all three samples were diluted 1:5 into mpox-negative saliva.

### Amplicon sequencing from saliva samples

To assess concordance of the primers and probes of the assays, we followed the sequencing protocol developed by Chen *et. al*. [20], after performing DNA extraction on the 30 known-positive samples collected into Spectrum SDNA-1000 tubes using the MagMAX™ Viral/Pathogen Nucleic Acid Isolation Kit (ThermoFisher Scientific, MA). After selecting specimens with Ct values <31 following PCR testing using the Logix Smart™ Mpox (2-Gene) RUO assay, we proceeded with sequencing seven saliva specimens using the Yale Center for Genomic Analysis’ Illumina MiSeq at a depth of 1.5 million reads, then followed the bioinformatics pipeline detailed by Chen *et al*. [20].

Next, we performed a reference-guided whole genome alignment using a reference sequence retrieved from the National Center for Biotechnology Information’s (NCBI) GenBank. We compared the nucleotide sequences of the primers and probes of the assays used in this study against the aligned sequences for concordance using Geneious Prime® v2023.1.1. We retrieved an additional 1,560 complete genomes from GenBank with collection dates from January 1^st^, 2022 through April 7^th^, 2023. The primers and probes of the assays were then compared across the new alignment which included a total of 1,484 sequences after sequence quality checks. Lineages were assigned using Nextcalde [21], according to Happi *et al*. [22].

### Statistical methods

Statistical analyses were performed using R v4.2.2 (2022-10-31) and data visualizations were produced using GraphPad Prism v9.4.1. We used a logistic regression model to see if there was a difference in detection as well as a one-way ANOVA to determine whether there was a difference in mean Ct values across assays.

To evaluate the impact of temperature over time, a linear model was used. An interaction term was used to evaluate the effect of time and temperature by sample (time*temperature). The ΔCt value represents the change in the Ct value from saliva under each condition (categorical). The reference group for time was zero, and the reference group for temperature was room temperature (∼19°C). Samples where no mpox virus DNA was detected were set to Ct = 45.00. P values of less than 0.05 were considered significant.

## RESULTS

### PCR assay performance

The limit of detection (LOD), determined using the synthetic mpox virus DNA (ATCC VR-3270SD) was 3 copies/µl for the Mirimus MPOX RT-PCR assay and 1 copy/µl for both CDC assays. The Logix Smart™ Mpox (2-Gene) RUO assay did not detect the synthetic DNA used so the LOD could not be determined. When testing DNA extracted from saliva collected in Spectrum SDNA-1000 collection devices, all assays performed comparably at detection (*p*>0.05). Mean Ct values generated did not differ across the assays (*p=*0.59).

### Extraction-free workflows for testing saliva for the detection of mpox virus

Detection of mpox virus in saliva was comparable across all three extraction-free workflows (**Supplementary Figure 1**). Due to limited sample size, it was not possible to statistically quantify the differences in performance of the workflows.

### Stability of detection of mpox virus in saliva

The detection of mpox virus remained relatively stable in raw saliva for 24-48 hours, with degradation notable at 72 hours (**Figure 1**). As compared to time zero, we observed an increase of 2.14 Ct, 2.55 Ct, and 4.72 Ct, at 24, 48, and 72 hours, respectively for Co-Diagnostic’s T3 target and an increase of 2.70 Ct, 3.05 Ct, and 4.99 Ct, respectively for the T4 target. When fitting the linear model with the interaction term, we found that at 40°C, temperature makes degradation worse over time. We observed similar results for the samples that were diluted 1:100 in negative saliva and tested with the Mirimus MPOX RT-PCR assay (see **Supplementary Figure 2**). When testing the stability of detection of naked mpox DNA spiked directly into raw saliva at room temperature, as compared to time zero, we observed a mean increase of <2 Ct at 24 hours and 4.2 Ct at 72 hours (see **Supplementary Table 4** for data).

**Figure 1.**
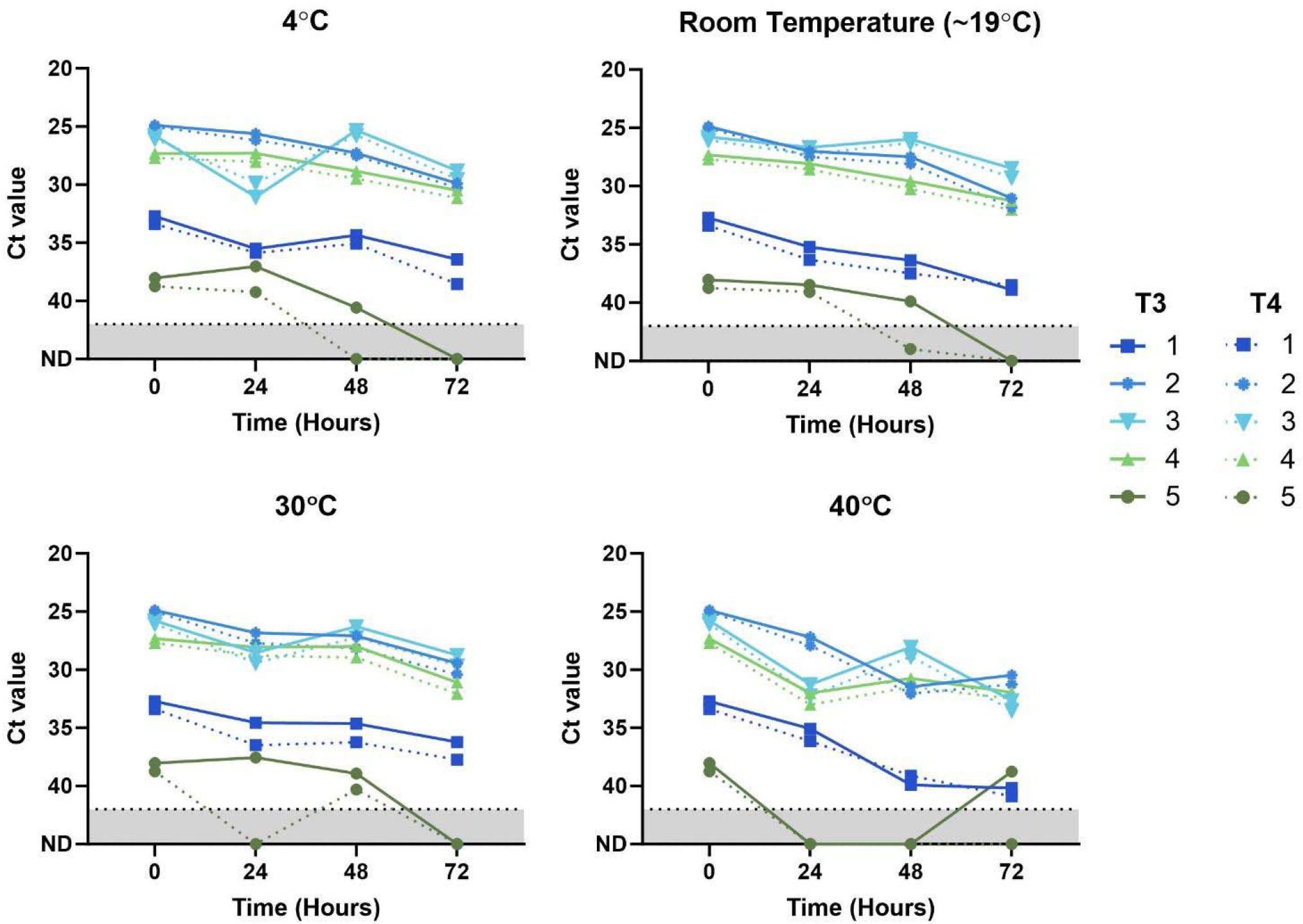
Stability of mpox virus detection in raw (unsupplemented) saliva after prolonged storage at different temperatures. Mpox-positive saliva samples were diluted 1:10 into raw, mpox-negative saliva. Prepared saliva lysates were tested by PCR using the Co-Diagnostics Logix Smart™ Mpox (2-Gene) RUO assay. The “T3” target cycle threshold (Ct) values are shown as solid lines, the “T4” target Ct values are shown as dotted lines. Results suggest that the detection of mpox virus remains relatively stable for approximately 24–48 hours across all storage conditions, with reduced stability in raw saliva at 40°C. While the low viral load in sample 5 resulted in its lack of detection at 24 and 48 hours of incubation at 40°C, the T4 target was detected at 72 hours, likely due to stochastic fluctuations at such low levels.

Detection of mpox virus in raw saliva also remained stable following the 56-hour incubation through temperatures and time periods simulating summer and winter shipping conditions (**Figure 2**), suggesting that saliva samples collected for testing for mpox may be sent through postal systems while maintaining sensitivity.

**Figure 2.**
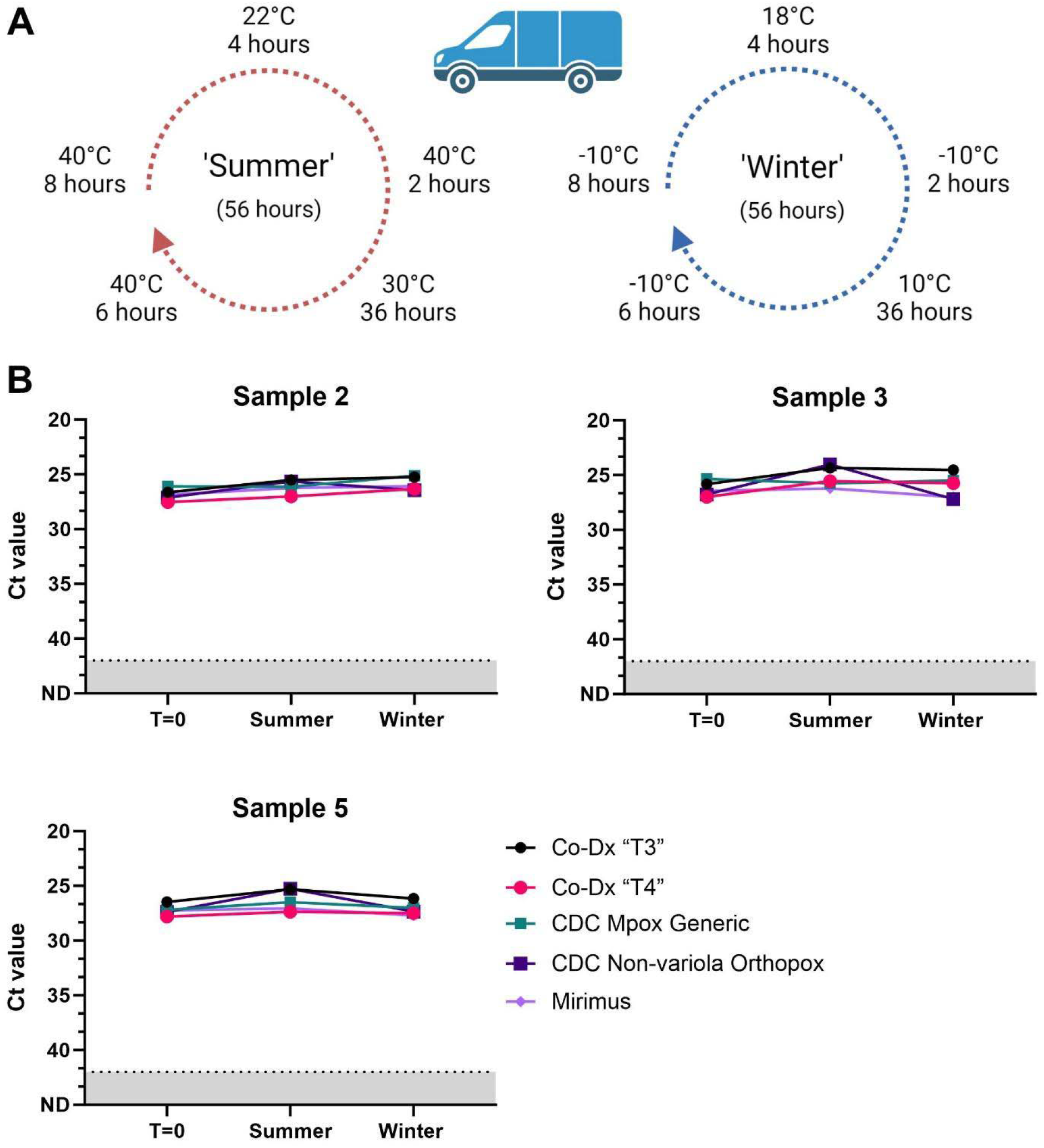
Detection of mpox virus in raw (unsupplemented) saliva remained stable after cycling through temperatures representing shipping conditions encountered in summer and winter. Three mpox-positive saliva samples were diluted 1:5 into raw mpox-negative saliva and (**A**) cycled through simulated summer and winter shipping profiles, recommended by the US Food and Drug Administration for the validation of remote sample collection. Following, saliva lysates were prepared following workflow one then tested by PCR using the Co-Diagnostics Logic Smart™ Mpox (2-Gene) RUO assay, CDC Monkeypox virus Generic Real-Time PCR assay, CDC Non-variola Orthopoxvirus Generic Real-Time PCR assay, and Mirimus MPOX RT-PCR assay. (**B**) Resulting cycle threshold (Ct) values were compared to the initial sample testing (T=0) and demonstrated stable detection of mpox, following sample incubation. Panel A created with biorender.com.

### Amplicon sequencing from saliva samples

We sequenced whole mpox virus genomes from ten saliva samples according to the amplicon-based sequencing protocol. We were able to sequence over 95% of the genome from seven of the ten samples (see Supplementary Table 3; GenBank Accession Numbers PP977060-PP977067). All of the samples sequenced clustered within clade IIb, falling within lineage B1. Three of the sequences were identified in sub lineages B1.2.1, B.1.4 and B.1.17.

After aligning the seven consensus sequences to the reference genome “NC_063383”, we visually examined the alignment of the forward primers, reverse primers, and probes across the different assays. We found two different mismatches with the CDC’s Monkeypox virus Generic RT-PCR Assay. The sequences had a single nucleotide substitution in the forward and reverse primers.

Importantly, the reference sequence collected in Nigeria in 2018, only had a mismatch in the forward primer; there were no nucleotide substitutions in the reverse primer or the probe. All primers and probes of the other assays were concordant with the generated sequence as well as the reference sequence. The discordant sequences and the primers/probes can be seen in Table 3. Furthermore, alignment with genomes retrieved from Genbank, confirmed that indeed the primers for the CDC’s Monkeypox virus Generic Real-Time PCR Test had several mismatches across the 1,484 genomes. Neither the Clade IIa Assay nor the CDC’s Non-variola Orthopoxvirus Generic Real-Time PCR Test primer and probe sets were affected.

**Table 3.**
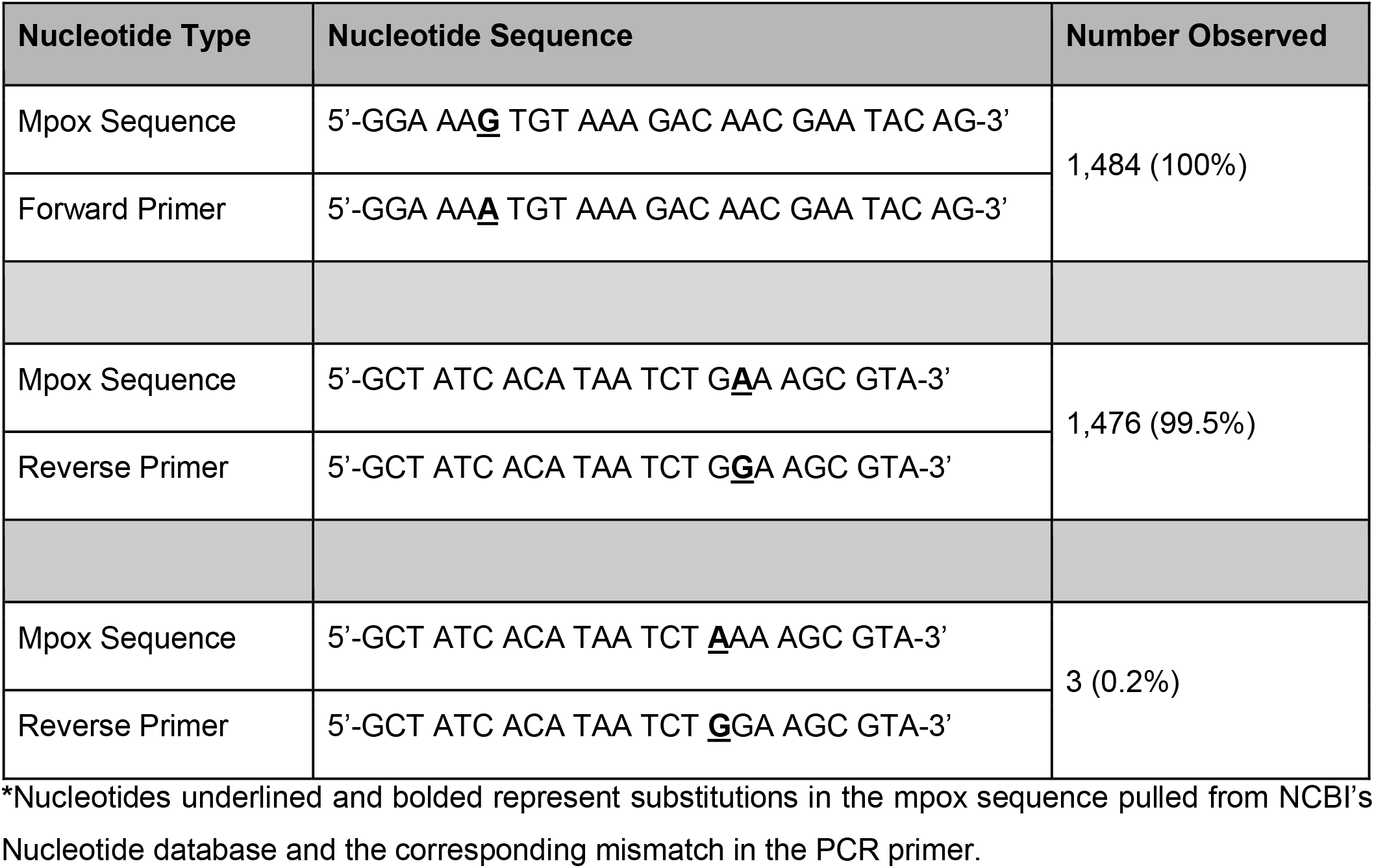
CDC Monkeypox virus Generic Real-Time PCR Test primer concordance with 1,483 sequences aligned to reference sequence ‘NC_063383’.

These findings mostly agree with the results reported by Wu *et al*. [23]. In our study, 100% of the 1,484 sequences had the A6G substitution in the forward primer, while 99.5% (n=1,476) sequences included in our alignment had the G17A substitution in the reverse primer. Additionally, we observed 3 sequences with a G16A substitution in the reverse primer – all of which were from the United States.

## DISCUSSION

Numerous studies since the 2022 outbreak of mpox have investigated the detection of mpox virus in saliva [5–9,12,24], reporting its high concordance with lesion swabs [8,24] and higher viral load (as approximated by Ct values) in saliva in comparison to other respiratory samples [9], and sometimes also lesion swabs [6,7]. With a geographically diverse network of clinical laboratories connected through the SalivaDirect FDA Emergency Use Authorization PCR test [25], we recognized its potential to greatly bolster the response to the US mpox outbreak. Leveraging previously published and commercially available assays to support the rapid adaptation of our streamlined PCR test for SARS-CoV-2 [26], we confirmed the comparable performance of five different PCR assays and incorporated them into our extraction-free protocol. Extraction-free workflows further reduce costs by minimizing laboratory overhead for sample processing, requiring less equipment and reagents, while facilitating timely results [9,26–28]. In countries where supply-chain infrastructure for test kit materials, reagents, and laboratory equipment may be strained, having a flexible platform may reduce barriers to testing while offering flexibility for fitting into existing laboratory regulations and/or processes.

To further support access to testing, we considered it essential to evaluate the stability of mpox detection in raw (unsupplemented) saliva to facilitate cold-chain-free options for both low-cost and remote and/or discrete sample collection, which holds potential as a means for frequent, follow-up testing in exposed individuals, before the appearance of lesions. Expanding upon the findings of Yinda *et al*., demonstrating stability of mpox virus spiked into saliva over the course of 20 days [29], our study demonstrates the stable detection of mpox virus DNA in raw saliva for 24-48 hours and under simulated seasonal shipping conditions. This suggests that, similar to recommendations made for variola virus in 1957 [30], samples transported in this window may not require stabilizing media, minimizing test-kit costs as only a collection tube is necessary; cold-chain transport of specimens is only necessary in settings where temperatures near 40°C. It is important to note that as the samples were diluted into mpox-negative saliva, the composition of the pooled negative saliva may have disrupted the integrity of the original sample, thereby negatively-impacting the stability of mpox virus DNA. Despite this, these findings support a possible low-cost solution for at-home sample collection which can provide a more discrete option for groups facing stigma.

Genomic sequencing of pathogens contributes to the understanding and monitoring of outbreaks and transmission chains. The ability to sequence from saliva further negates the requirement of swab-based samples which are typically considered necessary for this. Importantly, the multiple substitutions that we observed in the primers of the CDC’s assay highlight the ongoing need for routine genomic surveillance of emerging pathogens to ensure adequate diagnostic performance in the face of virus evolution. The mismatches identified by us and different mismatches reported by others [23,31] reaffirms the necessity of multiplexed assays and mpox-specific targets for reliable diagnosis.

We recognize that the small sample size remains a limitation of our preliminary investigation and that it emphasizes the need for prospective collection of multiple sample types during outbreaks of emerging pathogens to identify best diagnostic practices. While ≤5 clinical samples have been tested in this work, diluting these in negative saliva increased the number of contrived samples that we could validate across potential storage conditions and across different workflows and PCR assays. Consistent results across these settings is promising and as such, despite these limitations, the results of this study contribute to the expanding body of evidence that supports both extraction-free [9,27] and saliva-based methods for the detection of mpox virus.

Saliva is increasingly being accepted as a clinical sample type across the globe [32,33] and should be considered as a potential tool to aid in the ongoing global need for the timely diagnosis and surveillance of pathogens such as mpox virus. Our open-source PCR test demonstrates how low-cost options could be utilized to support this, particularly when implemented within a testing framework that can support a rapid and flexible outbreak response. However, further research is needed to assess the temporal dynamics of mpox virus in saliva, which will elucidate the window at which we can expect patients to become positive and/or infectious through saliva. As such, questions pertaining to asymptomatic transmission/prolonged viral shedding also remain and identifying these cases is important for halting ongoing human-to-human transmission. Combined, this understanding can aid us in prediction of when disease may be at its worst based on viral load. Therefore, we echo Coppens *et al*. for studies that prospectively screen close-contacts of confirmed cases to answer these questions [24]. The ongoing need for diagnostics development and sustainable surveillance methods is accentuated by case reports of reinfection, break-through infection following vaccination, and infection of those who have both prior infection and full vaccination [34–36].

## Data Availability

All data produced in the present study are available upon reasonable request to the authors.

## ACKNOWLEDGEMENTS

We thank the Grubaugh Laboratory at the Yale School of Public Health for providing guidance on the amplicon-sequencing protocol they developed – in particular Dr. Nathan D. Grubaugh, Nicholas F.G. Chen, and Dr. Chrispin Chaguza. We also would like to thank Mirimus Labs, Co-Diagnostics Inc., and Dr. Delphine Dean at Clemson University for providing us with the PCR assays used in this study. Research reported in this publication was supported by the National Institute of General Medical Sciences of the National Institutes of Health under

Award Number 1S10OD030363-01A1. We also thank Yale University’s Fund for Lesbian and Gay Studies for financially supporting this research.

## ROLE OF THE FUNDER

This work was supported by Yale University’s Fund for Lesbian and Gay Studies (FLAGS) and SalivaDirect, Inc..

## AUTHOR’S CONTRIBUTIONS

ALW conceived and designed the study. OMA, DY-C and ALW managed the study. TZ, MB, SS and JP managed sample collection. RJT, OMA, DY-C and KF were responsible for and performed the assays. RJT, OMA and ALW performed the analyses and interpreted the data. RJT, CB, SAS, OMA and ALW drafted the manuscript. All authors amended and commented on the final manuscript.

## DISCLOSURES

ALW has received consulting and/or advisory board fees from Pfizer, Merck, Diasorin, PPS Health, Primary Health, Co-Diagnostics, and Global Diagnostic Systems for work unrelated to this project, and is Principal Investigator on research grants from Pfizer, Merck, and NIH RADx UP to Yale University and from NIH RADx, Balvi.io and Shield T3 to SalivaDirect, Inc.. All other co-authors declare no potential conflict of interest.

## Notes

### Author Declarations

This study was conducted with Institutional Review Board approval from Yale Human Research Protection Program (Protocol ID. 2000033293), which allowed for the use of remnant clinical samples and was considered as non-human subjects research. No personal identifiable information was used for this study.

### Summary of Updates

Additional supporting literature has been added, additional DNA stability studies, GenBank Accession Numbers of submitted sequences.

